# Comparative Application of the Fluctuation Test to the data of Morbidity by COVID-19 in United States of America, United Kingdom, Taiwan and China 2020-2023

**DOI:** 10.1101/2024.04.06.24305422

**Authors:** Anyi Yannire López-Ramírez, Hilda Cristina Grassi, Efrén de Jesús Andrades, Jesús Enrique Andrades-Grassi

**Affiliations:** Graduate Student of Master Program, PQM, Facultad de Farmacia y Bioanálisis, Universidad de Los Andes (ULA), Mérida, Venezuela; Sección de Biotecnología, Instituto de Investigaciones, Facultad de Farmacia y Bioanálisis, Universidad de Los Andes (ULA), Mérida, Venezuela; Departamento de Ordenación de Cuencas, Facultad de Ciencias Forestales y Ambientales, Universidad de Los Andes (ULA), Mérida, Venezuela.

**Keywords:** Fluctuation Test, COVID-19, Morbidity, SARS CoV-2, USA, UK, Taiwan, China

## Abstract

In this work the Luria and Delbruck Fluctuation Test was comparatively applied to the data of Morbidity by COVID-19 in the United States of America (USA), United Kingdom (UK), Taiwan and China from 2020 to 2023. Three types of data were used: es.statista.com, datosmacro.expansion.com and larepublica.co without modification, but trying to avoid and justify the anomalies and inconsistencies observed. The methods originally used to establish the interactions of two populations were evaluated: the viral population with that of its host and the drift of both organisms. Only the interactive fluctuations of the weekly Variance of daily increase of Cases (Morbidity) were studied. The results showed that the Fluctuation Test is applicable to the selected data from USA, UK, Taiwan and China and other data from several countries used as controls. The study was separated into two approaches: First, comparison of the total or partial logarithmic profile of fluctuations of Variance of Cases (Morbidity) of USA, UK, Taiwan and China. Second, comparison of the values of the first fluctuation of Variance of Cases (Morbidity) in the boreal winter of 2020 for USA, UK, Taiwan, China and several countries used as controls. The results obtained for Morbidity demonstrate that USA and UK present a similar bimodal profile. China shows an inverted profile and Taiwan shows an intermediate profile between both tendencies. However, it was possible to detect some anomalies and uncertainties that were possibly derived from inconsistencies in the original data. Only USA shows a value of the first fluctuation comparable to the order of magnitude of the value of the first fluctuation of the Variance of Cases of China, in the northern winter of 2020. In the First Approach USA, UK and China had two important fluctuations: the first in the northern winter of 2020 before week 16 and the second at the beginning of northern winter of 2022, more than 100 weeks later. Taiwan showed only the latter. This latest fluctuation coincides with two events: the possible achievement of herd immunity and the emergence of Omicron variant. In this work we have evaluated whether this coincidence is casual or causal. The results obtained in the Second Approach aim to confirm the hypothesis of the animal origin of the first variant of SARS CoV-2.

## Introduction

SARS CoV-2 virus is the etiological agent for COVID-19 outbreak in China **[1]**. This first variant that emerged in a food market in Wuhan, Hubei province in late 2019 produced several derived variants **[2]** as this emerging infection spread worldwide **[3]**. Due to the continue global spread of COVID-19 and the lack of specific medicine, vaccination including compromised patients boosting has been the main mean to prevent and control COVID-19 in many countries **[4]**. The World Health Organization (WHO) on March 11, 2020, has declared the novel Coronavirus (SARS CoV-2) outbreak a global pandemic (COVID-19) **[5]**.

In this work we tried to adapt the Luria and Delbruck Fluctuation Test **[6]** to the data of USA, UK, Taiwan, China and other countries used as controls, reported on cases from three sources of information to verify its applicability, to show and analyze the results and the information that can be produced and to contribute with some aspects such as the Origin of the first variant of the SARS virus CoV-2 **[7]**, the role of Omicron variant **[8], [9]** and the possible achievement of herd immunity.

## Methods

The Luria and Delbruck Fluctuation Test was adapted according to the method originally used to establish the interaction of two types of populations: The Viral population (SARS CoV-2) and the Host population (human beings from USA, UK, Taiwan, China and other countries used as controls). Countries studied in alphabetical order: Australia (Aus), Brazil (Bra), Canada (Can), China (Chi), France (Fra), Germany (Ger), Iceland (Ice), India (Ind), Italy (Ita), Japan (Jap), Mexico (Mex), South Africa (SA), South Korea (SK), Spain (Spa), Taiwan (Tai), United Kingdom (UK), United States of America (USA).

The veracity of the data was not examined. If in said data there are involuntary or voluntary errors or misconduct, the results, analysis and conclusions of this work will reflect them. Three types of data were used: es.statista.com; datosmacro.expansion.com and larepublica.co without modification, but trying to avoid and justify the anomalies and inconsistencies observed. We proceeded to download the data of the number of confirmed cases and their daily increase (Morbidity). This data was tabulated in several Excel tables to facilitate its visualization and the calculations of the average and weekly variance of daily increase in Morbidity in the years 2020, 2021, 2022 and 2023. Changing the start and end of the days of some of the weeks under study was tested for different data sources, different periods and different countries, to verify whether this shift would produce important changes in the results. The location of the fluctuations was not affected. Furthermore, the values found were of the same order and similar. A discontinuous graphic system was designed to avoid data with anomalies and inconsistencies and to allow the treatment of different data sources, in different periods. Moreover, the period in which the full vaccination reached more than 75 per cent and the herd immunity was supposedly hit in China, was considered to separate the study of China in two periods. Like China **[10]**, in this study USA, UK, and Taiwan have an important Fluctuation in the northern winter of 2022.Two events appeared simultaneously along with this Fluctuation: the possible achievement of herd immunity and the emergence of Omicron variant. This coincidence could be by chance or causal. Following the Luria and Delbruck ideas **[6]** in this work the possible causality has been hypothetically explained as the Natural Selection of new variants of SARS CoV-2 that appeared spontaneously. The selective pressure on the SARS CoV-2 population could be due to the fact that the human population reached levels of immune response and treatment against COVID-19 infection that tended to control and stop the development of the virus. In this work, the term immune response includes all systems and reactions to infection with SARS CoV-2. Only fluctuations in the Variance of Cases (Morbidity) are reported. The Figures and the Table express the logarithm of Variance in relation to the Weeks of each year. A maximum total number of Weeks of 52 is considered in each year. Calculation of the Logarithm of the Variance of Cases is done only for reasons of presentation and visualization of results. This new value does not represent an additional parametric study. Zero and negative values would be explained due to the presence of anomalies in the Data.

This study does not belong to formal Statistic Sciences. Instead it is an exploratory and comparative application of the Fluctuation Test of Luria and Delbruck that evaluates the resulting logarithmic profile from the calculation of Weekly Variance (as VAR.P) of official data reported for COVID-19 cases for USA, UK, Taiwan, China and other countries used as controls. In this study, the calculation of Weekly Variance (as VAR.P) of cases is applied to verify the appearance or not of Fluctuations that would be expressed as a consequence of some events such as quarantine and strict confinement, the appearance of new variants of SARS CoV-2 and vaccination campaign. The calculation of Weekly Variance of the daily increases in cases from COVID-19 official reported data, were carried out according to VAR.P (number 1,{number 2} …) in the Excel spreadsheet.

## Results and Discussion

### First Approach

es.statista.com, datosmacro.expansion.com, larepublica.co.

Figure 1 shows the weekly logarithmic profile of variance of cases of COVID-19 between the beginning of 2020 and the beginning of 2023 in USA. The profile is Bimodal with a Biphasic polynomial tendency, just like that shown for UK in Figure 2. The first and second phases last approximately one year (from 50 to 60 weeks each one), with the beginning of each phase coinciding with the boreal winter.

**Figure 1.**
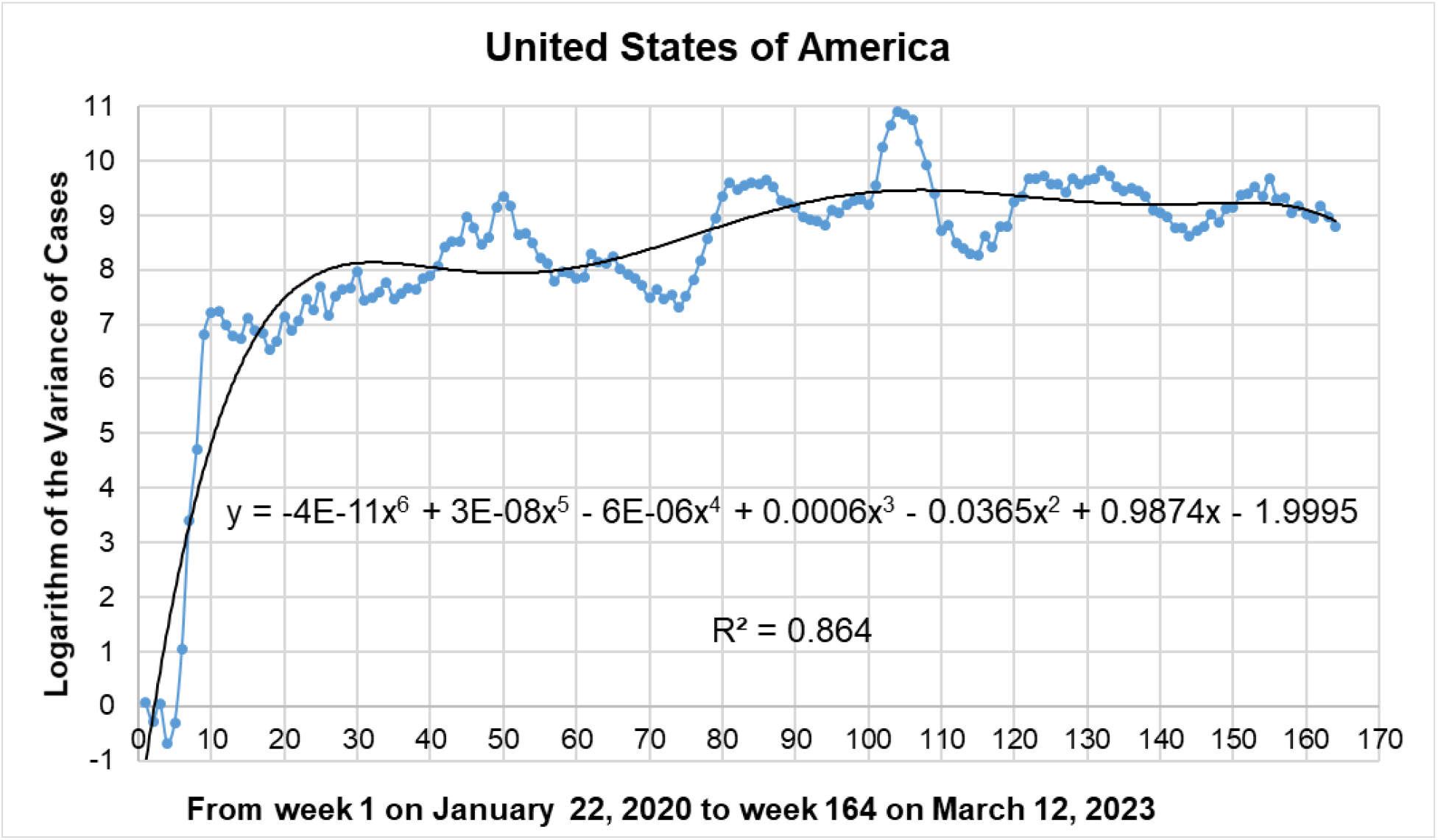
Logarithm of the variance of cases of USA based on 164 weeks, from January 22, 2020 to March 12, 2023.

**Figure 2.**
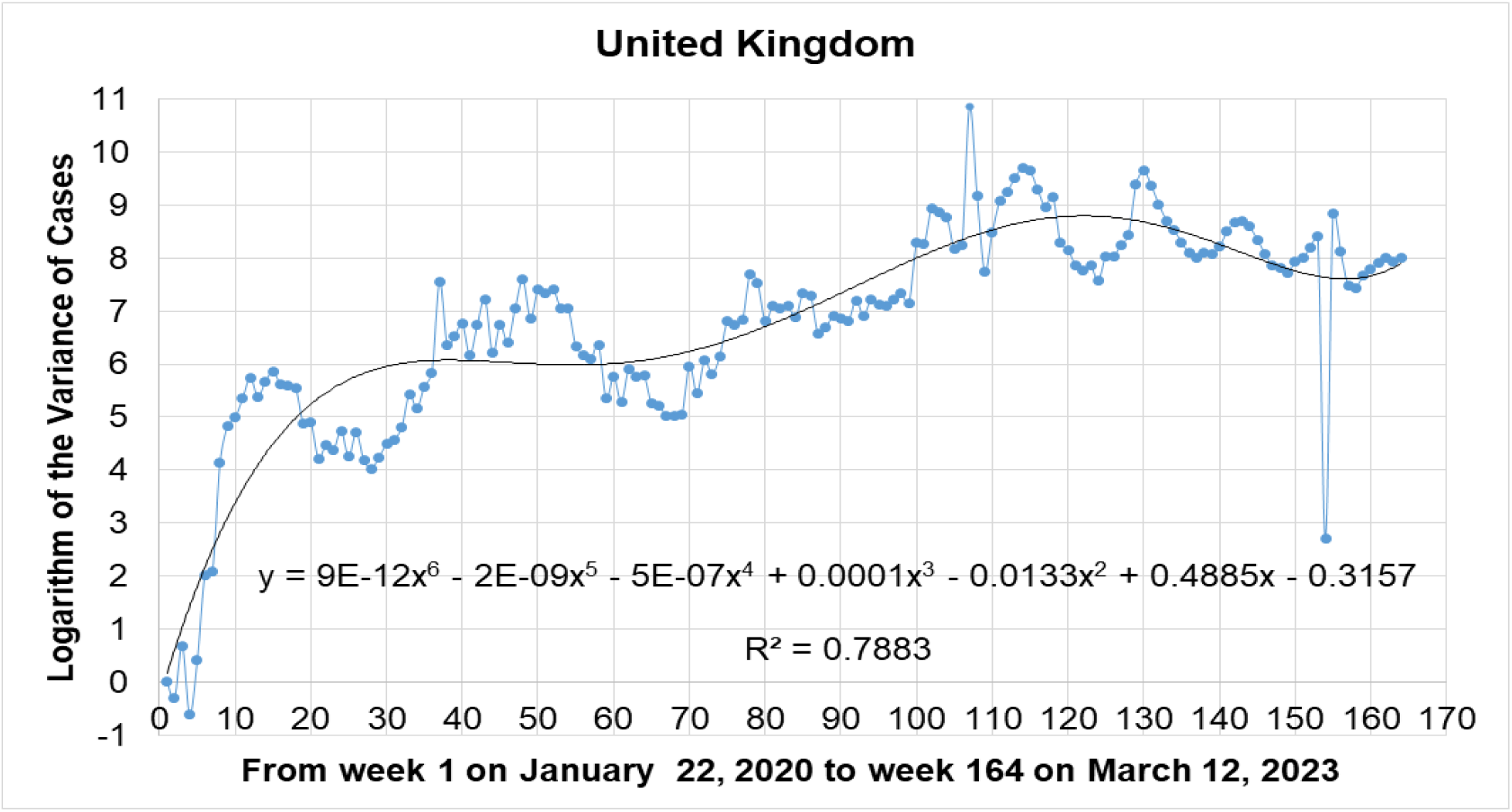
Logarithm of the variance of cases of UK based on 164 weeks, from January 22, 2020 to March 12, 2023.

In the case of the First Period **[10]** in China (see Figure 3) the profile is inverted and symmetric, with two “back to back” fluctuations and an intermediate “plateau” between the beginning of 2020 and the beginning 2022, confirming the effect of the Zero COVID-19 Policy **[11]** applied in that country. In the Second Period (end of 2021 to beginning of 2023, see **[10]**) of China, from the end of 2021 to the beginning of 2022, Figure 4 (datosmacro.expansion.com, larepublica.co) shows the same final fluctuation (week 110) observed in Figure 3 (es.statista.com).

**Figure 3.**
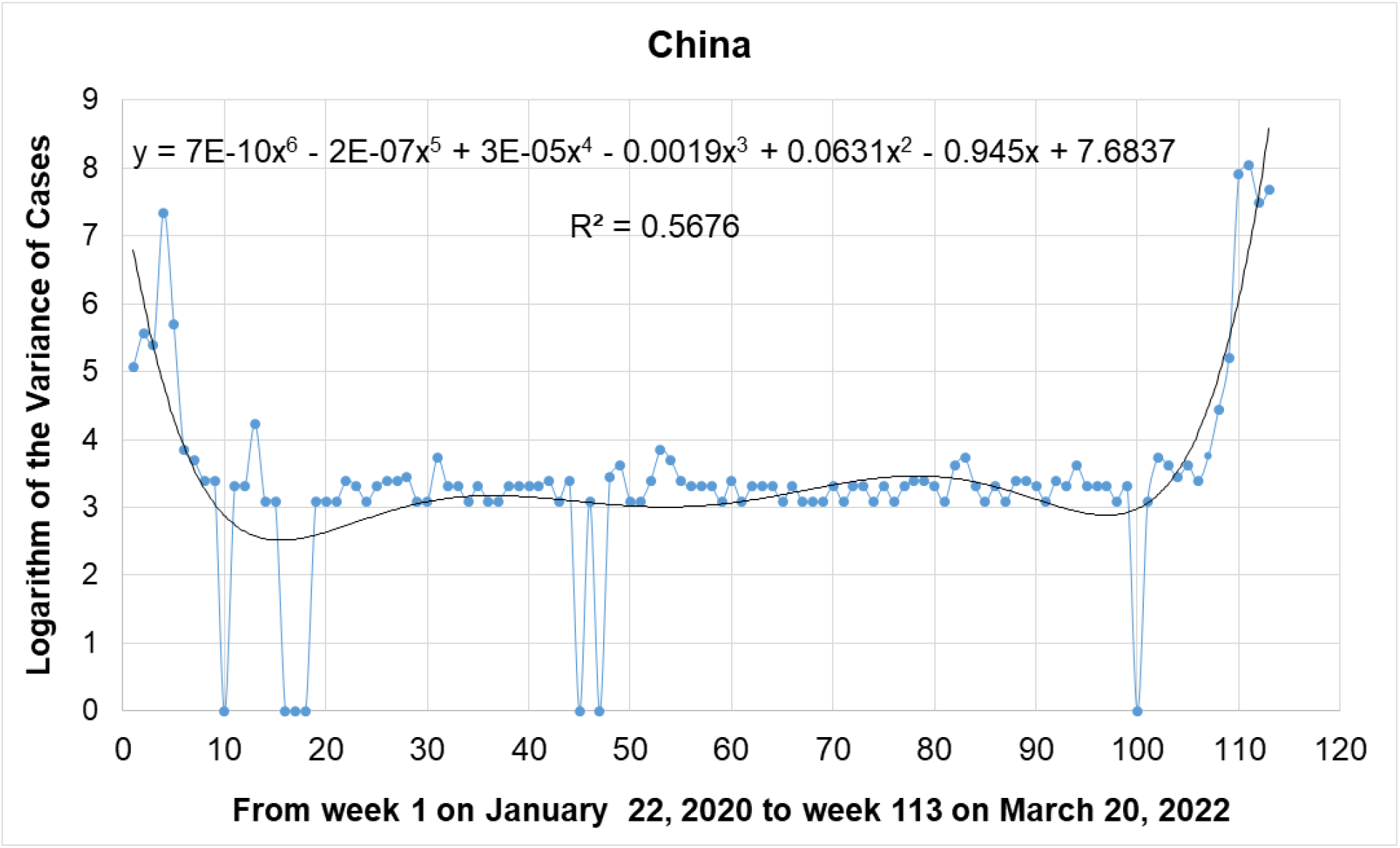
Logarithm of the variance of cases of China based on 113 weeks, from January 22, 2020 to March 20, 2022.

**Figure 4.**
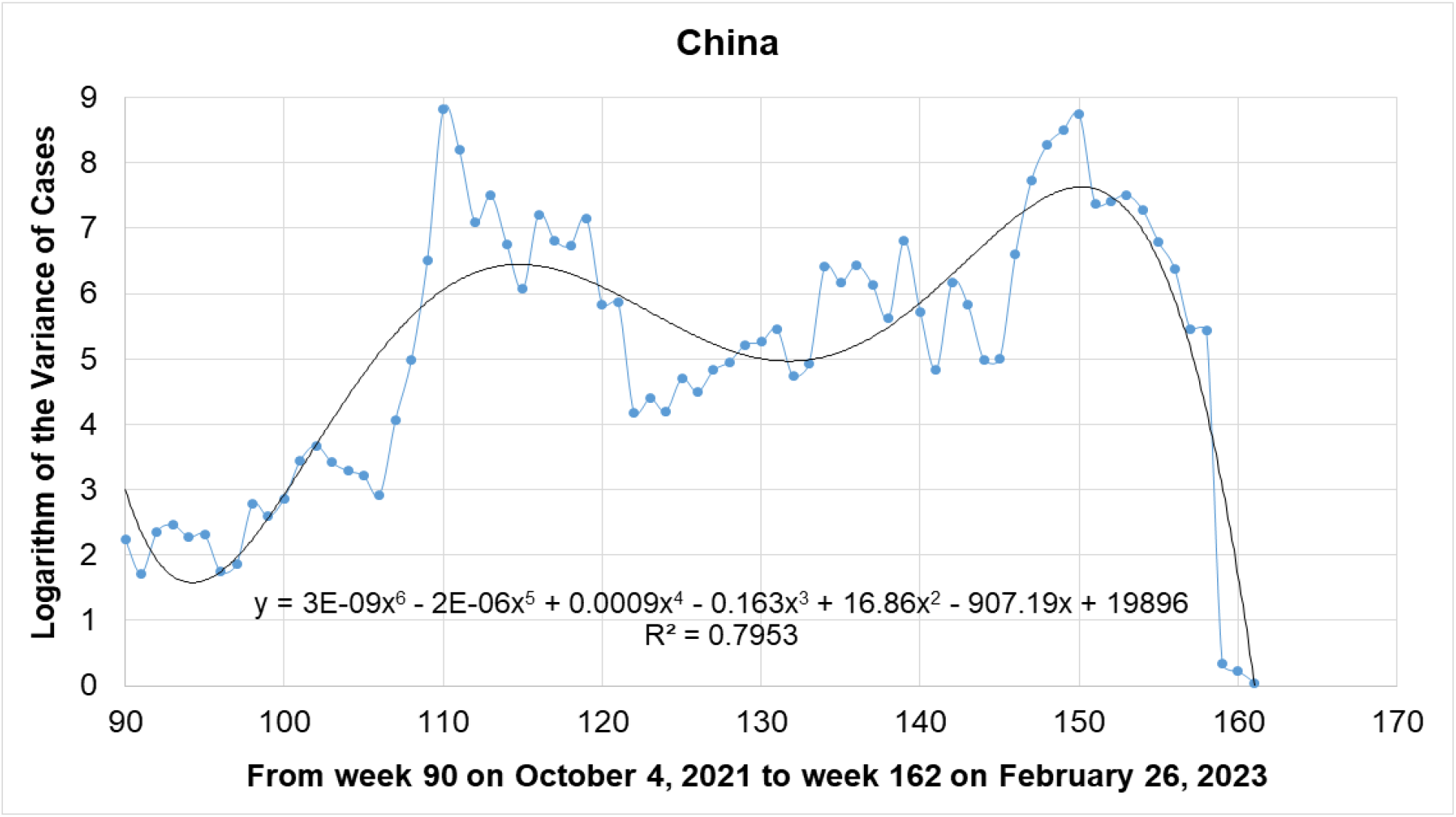
Logarithm of the variance of cases of China from October 4, 2021 (week 90) to February 26, 2023 (week 162).

Figure 5 shows the profile obtained for Taiwan between the years 2020 and 2023: It does not present the first important fluctuation of northern winter 2020. However, Taiwan and all other countries show an important fluctuation at the beginning of the northern winter 2022. This fluctuation coincides with the appearance of the Omicron variant and possibly with the herd immunity.

**Figure 5.**
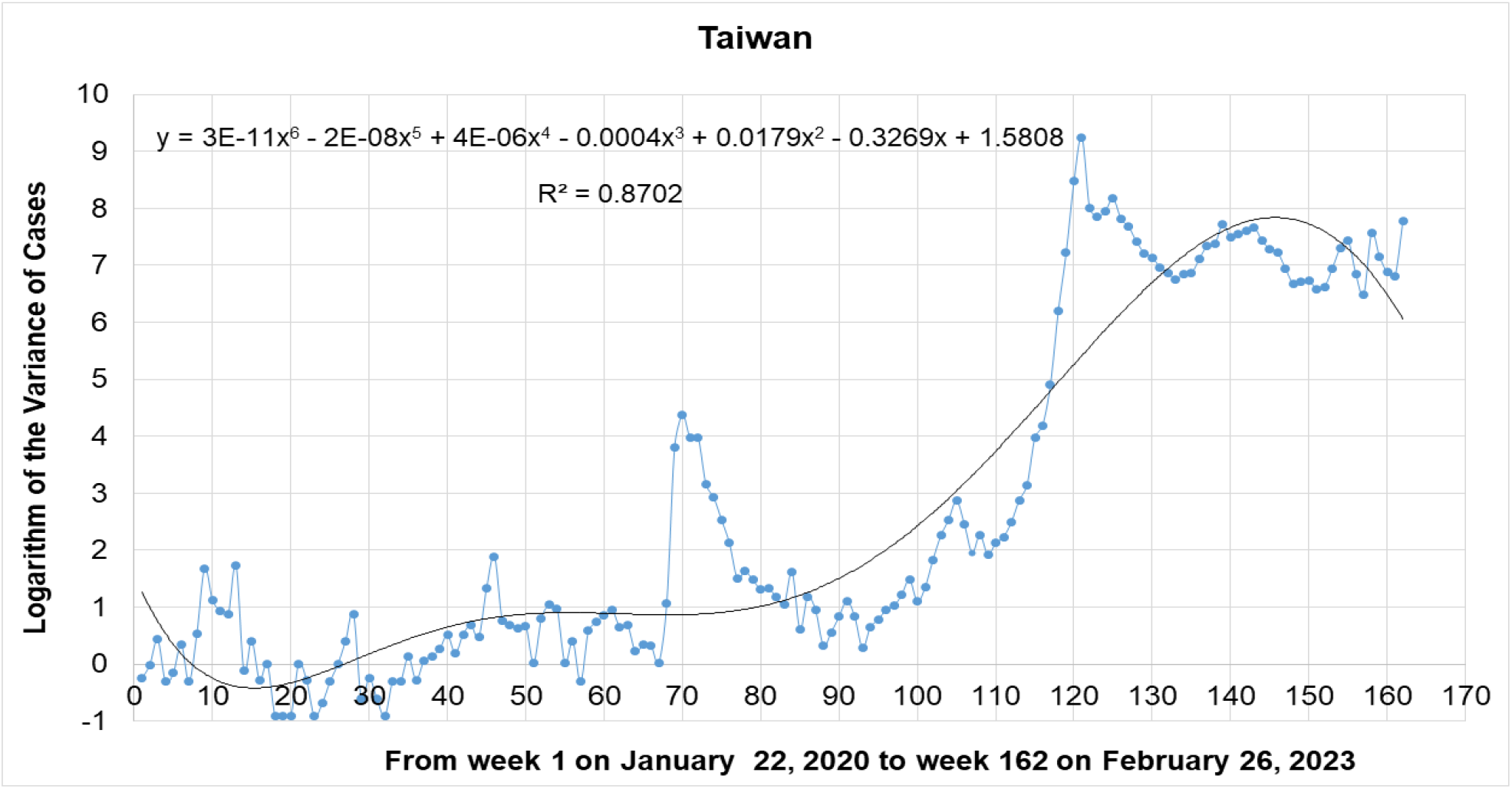
Logarithm of the variance of cases of Taiwan based on 162 weeks, from January 22, 2020 to February 26, 2023.

### **Second Approach:** larepublica.co.

An attempt was made to compare the values of the logarithm of variance of the first fluctuation of the boreal winter of 2020 in seventeen countries. These results are shown and analyzed in this section.

The period around March 16 and 23, 2020, was the most important for the start of the Pandemic in the countries studied in this work (see Table 1). USA presents a value in the first fluctuation of the same order of magnitude as China. The five countries that follow are from Europe and have values of a much lower order of magnitude. Only China and USA present values of the logarithm of variance greater than 7, Spain and Italy follow them with values greater than 6. Ten countries show values of logarithm of variance less than 6 and greater than 4. Furthermore, the profile of the graph (see Figure 6) presents an inflection in that segment.

**Table 1.** All countries: Comparison of the values of the logarithm of the variance of cases of the first fluctuation of the boreal winter of 2020.

| # | Country | Week | Weekly Period of 2020 | Logarithm of the Variance of Cases |
| --- | --- | --- | --- | --- |
| 1 | Chi | 4 | February 10 to February 16 | 7.35 |
| 2 | USA | 11 | March 30 to April 05 | 7.25 |
| 3 | Spa | 10 | March 23 to March 29 | 6.31 |
| 4 | Ita | 9 | March 16 to March 22 | 6.14 |
| 5 | Ger | 10 | March 23 to March 29 | 5.85 |
| 6 | UK | 12 | April 06 to April 12 | 5.75 |
| 7 | Fra | 8 | March 09 to March 15 | 5.31 |
| 8 | Mex | 14 | April 20 to April 26 | 4.71 |
| 9 | SK | 6 | February 24 to March 01 | 4.68 |
| 10 | Ind | 11 | March 30 to April 05 | 4.65 |
| 11 | Can | 11 | March 30 to April 05 | 4.43 |
| 12 | Bra | 9 | March 16 to March 22 | 4.37 |
| 13 | Jap | 12 | April 06 to April 12 | 4.34 |
| 14 | Aus | 9 | March 16 to March 22 | 4.28 |
| 15 | SA | 10 | March 23 to March 29 | 3.69 |
| 16 | Ice | 9 | March 16 to March 22 | 2.92 |
| 17 | Tai | 9 | March 16 to March 22 | 1.68 |

**Figure 6:**
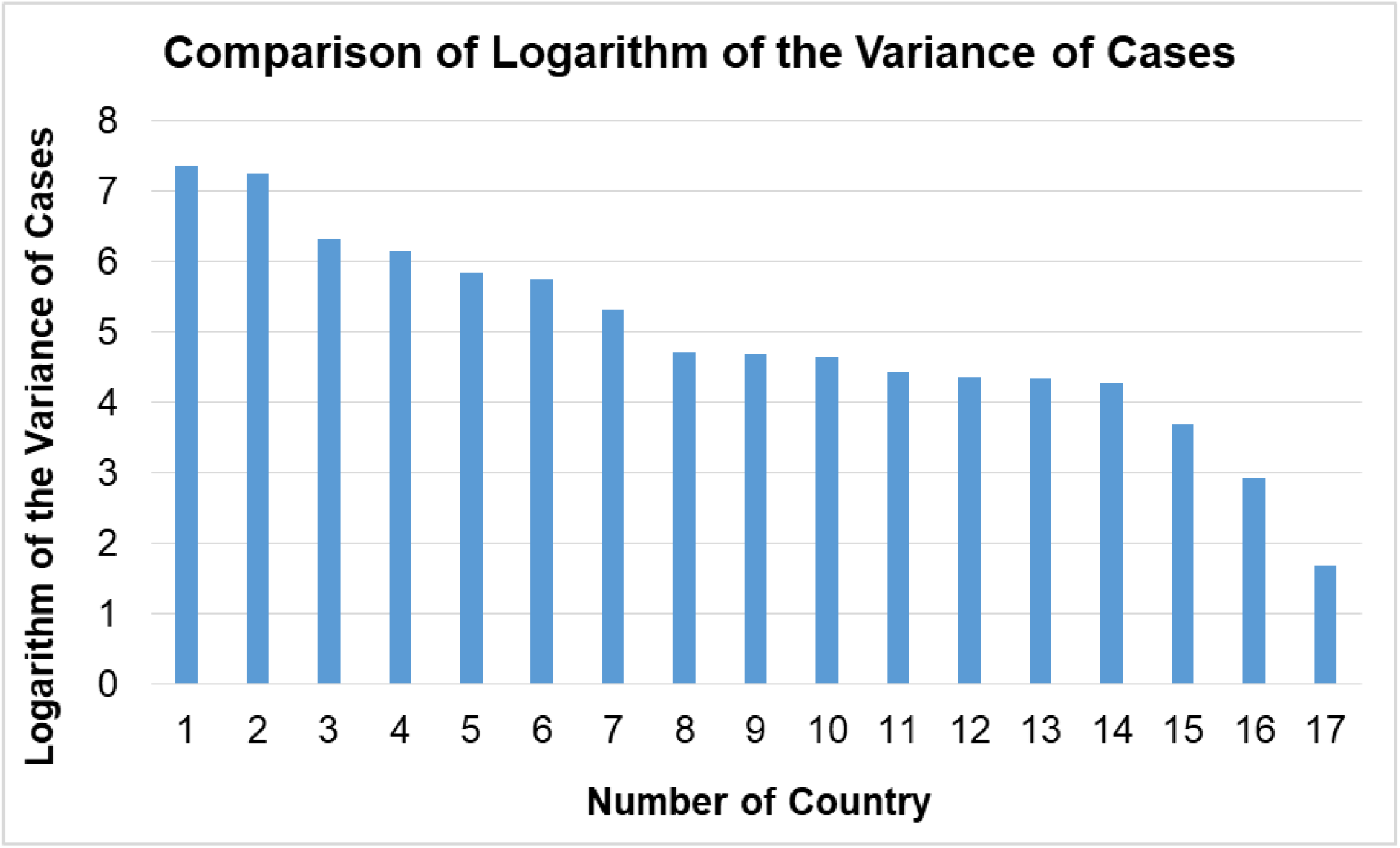
All countries: Comparison of logarithm of the variance of cases of the first fluctuation of the boreal winter of 2020.

These results would seem to coincide with the Tourism Gross Domestic Product (GDP) profile in the regions studied **[12]**.

## Conclusions

The data used in this work absolutely does not include the participation or opinion of the authors. It is declared that due to this, results, analysis and conclusions depend strictly on the veracity of said data.

The methods originally used to establish the interactions of two populations were evaluated: the viral population with that of its host and the drift of both organisms. The results showed that the Fluctuation Test is applicable to the selected data from USA, UK, Taiwan, China and other countries.

The results obtained for Morbidity demonstrate that USA and UK present a similar bimodal profile. China shows an inverted profile and Taiwan shows an intermediate profile between both tendencies.

In the First Approach USA, UK and China had two important fluctuations: the first in the northern winter of 2020 before week 16 and the second at the beginning of northern winter of 2022, more than 100 weeks later. Taiwan showed only the latter. This latest fluctuation coincides with two events: the possible achievement of herd immunity and the emergence of Omicron variant. In this work we have evaluated whether this coincidence is random or causal: following the ideas of Luria and Delbruck **[6]** it is concluded that the possible Darwinian causality may be explained as the Natural Selection of new variants of SARS CoV-2 that appeared randomly and spontaneously. The results obtained in the Second Approach aim to confirm the hypothesis of the animal origin of the first variant of SARS CoV-2: only USA shown a value of the first fluctuation comparable to the order of magnitude of the value of the first fluctuation of the Variance of Cases of China, in the northern winter of 2020.

## Data Availability

All data produced in the present study are available upon reasonable request to the authors

## Acknowledgements

Research Project: Application of Fluctuation Test and other Tools to Human Population Viral Infections (2020). Project Manager, EDJA.

The authors acknowledge to Maria Alejandra Fernandez-Ramirez who collected part of the data of China.

This work was financed by the Authors. The Authors declares no Competing or Conflict of interests.

## Author Contribution

HCG, EDJA and JEA-G: Conceived and designed the analysis. Contibuted analysis tools.

AYL-R and EDJA: Collected the data and performed the analysis.

AYL-R and EDJA: Drafting the manuscript and wrote final version.

## Bibliography

[1] Gorbalenya, A.E.; Baker, S.C.; Baric, R.S.; de Groot, R.J.; Drosten C.; Gulyaeva, A.A.;Haagmans, B.L.; Lauber C.; Leontovich, A.M.; Nueman, B.W.; Penzar D.: Perlman S.; Poon L.L.M.; Samborskiy, D.V.; Sioorov, I.A.; Sola I. and Ziebuhr, J. (2020). The species SARS-related-Coronavirus: Classifying 2019-nCoV and naming it SARS-CoV-2. Nat. Microbiol.5: 536–544. 10.1038/s41564-020-0695-z

[2] Pan, Y.; Wang, L.; Feng, Z.; Li, F.; Shen, Y.; Zhang, D.; Liu, W.J.; Gao, G.F. and Wang, Q. (2023). Characterisation of SARS-CoV-2 variants in Beijing during 2022: an epidemiological and phylogenetic analysis. Lancet. 401: 664–672. 10.1016/S0140-6736(23)00129-0

[3] Andrades-Grassi, J.E.; Cuesta-Herrera, L.; Bianchi-Perez, G.; Grassi, H.C.; Lopez-Hernandez, J.Y. and Torres-Mantilla, H. (2021). Spatial analysis of risk of morbidity and mortality by COVID-19 in Europe and the Mediterranean in the year 2020. Cuadernos Geograficos. 60: (1), 279–294.10.30827/cuadgeo.v60i1.15492

[4] Ripabelli, G., Sammarco, M.L., Rezza, G., D’Amico, A., De Dona, R., Lafigliola, M., Parente, A., Samprati, N., Santagata, A., Adesso, C., Natale, A., Di Palma, M.A., Cannizzaro, F., Dentizzi, C., Stefanelli, P., Tamburro, M. (2022). A SARS CoV-2 Outbreak among Nursing Home Residents Vaccinated with a Booster Dose of mRNA COVID-19 Vaccine. J. Community Health. 47: 598–603. Published on line: March 25, 2022. DOI: 10.1007/s10900-022-01082-8

[5] Cucinotta, D. and Vanelli, M. (2020). Who declares COVID-19 a Pandemic. Acta Bomed. Published on line (Internet) on March 19, 2020; 91 (1):157–160. Available from: https://www.mattioli1885journals.com/index.php/actabiomedica/article/view/9397 DOI: 10.23750/abm.v91i1.9397 PMID:32191675

[6] Luria, S.E. and Delbruck, M. (1943). Mutation in bacteria from virus sensitivity to virus resistance. Genetics, 28: 491–511.

[7] Alwine, J.C.; Casadevall, A.; Enquist, L.W.; Goodrum, F.D. and Imperiale, M.J. (2023). Editorial: A critical analysis of the evidence for the SARS-CoV-2 Origin Hypothesis. mBio, 14, (2): 1–7. 10.1128/mbio.00583-23

[8] Cai, J.; Deng, X.; Yang, J.; Sun, K.; Liu, H.; Chen, Z.; Peng, C.; Cheng, X.; Wu, Q.; Zou, J.; Sun, R.; Zheng, W.; Zhao, Z.; Lu, W.; Liang, Y.; Zhou, X.; Ajelli, M. and Yu, H. (2022). Modeling transmission of SARS-CoV-2 Omicron in China. Nat. Medicine, 28: 1468–1475. https://www.nature.com/naturemedicine 10.1038/s41591-022-01855-7

[9] Ren, S.Y., Wang, W.B., Gao, R.D. and Zhou, A.M. (2022). Omicron variant (B.1.1.529) of SARS CoV-2: Mutation, infectivity, transmission, and vaccine resistance. World J. Clin. Cases; 10 (1): 1–11. URL:https://www.wjgnet.com/2307-8960/full/v10/i1/1.htm DOI:10.12998/wjcc.v10.i1.1

[10] Lopez-Ramirez, A.Y., Fernandez-Ramirez, M.A., Grassi, H.C., Andrades, E.D.J., Andrades-Grassi, J.E. (2023) Application of the Fluctuation Test to the data of Morbidity and Mortality by COVID-19 in China 2020-2023. medRxiv 2023.11.12.23298174; 10.1101/

[11] Burki, T. (2023). Editorial published Online December 16, 2022: Moving away from Zero COVID in China. https://www.thelancet.com/respiratory.11. 10.1016/S2213-2600(22)00508-2

[12] Statista (2024). Direct contribution of Tourism to world Gross Domestic Product (GDP) by Regions in the year 2019. Published by Statista Research Departament on January 30, 2024, in Spanish https://es.statista.com/estadisticas/601454/

